# The Role of Coronary Artery Calcification in Metal-Related Cardiovascular Disease

**DOI:** 10.1101/2025.08.11.25333464

**Authors:** Arce Domingo-Relloso, Katlyn E McGraw, Irene Martinez-Morata, Yuchen Zhang, Kathrin Schilling, Ronald A. Glabonjat, Ziqing Wang, Kiros Berhane, Brent A. Coull, Marta Galvez-Fernandez, Miranda R. Jones, Wendy S. Post, Joel Kaufman, Tiffany R. Sanchez, Maria Tellez-Plaza, Graham R. Barr, Steven Shea, Ana Navas-Acien, Linda Valeri

## Abstract

**Background:** Metals are associated with cardiovascular disease (CVD), but the underlying pathways remain largely unclear. We evaluated the potential intermediate role of coronary artery calcification (CAC) trajectory on the association between urinary metals and incident CVD, accounting for competing risks by death from other causes.

**Methods:** We used data from 6,527 participants of the Multi-Ethnic Study of Atherosclerosis (MESA). CAC was measured longitudinally using the spatially weighted calcium score in five exams, starting in 2000. Participants were followed for CVD events through 2019. Cadmium, cobalt, copper, uranium, tungsten, and zinc were measured in urine at the baseline visit (2,000-2,002). We used a causal inference algorithm with a path-specific effects approach for longitudinal mediation analysis to evaluate the intermediate role of CAC on the association between metals and incident CVD.

**Results:** During follow-up, 1,140 participants had a CVD event and 1,147 died. The association with incident CVD mediated through the CAC trajectory was statistically significant for all metals except for uranium. The number of CVD cases (95% CI) per 100,000 person-years attributable to an interquartile range (IQR) increase in metal levels through the longitudinal trajectory of CAC was 39 (16, 66) for cadmium, 17 (0, 37) for cobalt, 20 (3, 38) for copper, 21 (3, 40) for tungsten, and 42 (24, 64) for zinc. The results were substantially attenuated for copper and zinc after adjusting for CVD risk factors, but not for the other metals.

**Conclusions:** This study supports that part of the association between urinary metals and CVD is attributable to changes in CAC over time. In particular, half of the association between urinary cadmium and CVD might be mediated by longitudinal changes in CAC. This study could inform strategies for early detection and prevention of CVD based on urinary metal levels.

## Introduction

Physicians are increasingly considering the importance of incorporating measurements of environmental exposures to clinical practice.^1^ Metals are cardiovascular disease (CVD) risk factors, as supported by mounting epidemiologic and mechanistic evidence.^2–5^ Exposure to metals might accelerate the development of atherosclerosis, including the accumulation of variable amounts of calcium in the coronary arteries (i.e.,coronary artery calcification (CAC)).^6^ CAC is traditionally measured non-invasively through cardiac computed tomography (CT), and can be quantified using a novel semi-automated, threshold-free method called spatially weighted calcium score (CAC-SW.^7,8^

CAC-SW levels are highly predictive of coronary heart disease (CHD) events.^9,10^ In a previous study, we found that non-essential (cadmium, tungsten, uranium) and essential (cobalt, copper, zinc) metals are longitudinally associated with CAC in the Multi-Ethnic Study of Atherosclerosis (MESA).^11^ We also found that the same metals are prospectively associated with clinical cardiovascular events including incident CVD and CHD.^12^ The amount of the association between different metals and incident CVD that might be explained by changes in CAC over time, however, is unknown.

Mediation analysis methods do not generally incorporate longitudinal measures of the potential mediator, and thus fail to incorporate changes over time preceding the clinical outcome. In addition, mediation analysis generally ignores competing risks by death or reports disease risk either reconstructing a population in which no one dies or considering death as a censoring event.^13^ This leads to biased estimates, as the competing event (i.e., death) makes it impossible for the event of interest (e.g., CVD) to happen, and information on whether participants would have developed the outcome is missing for those who die during the follow-up.

The MESA includes more than 6,000 participants from multiple centers across the US with different ethnicities and a long follow-up of almost two decades with repeated measures of CAC over time. These characteristics provide a unique opportunity to study the potential intermediate role of the longitudinal changes of CAC on the association between urinary metals and CVD risk. In this work, we used a novel approach to quantify the mediating role of CAC, measured longitudinally at five time points using CAC-SW, on the association between baseline urinary metal biomarkers and clinical CVD events. Our algorithm accounts for competing risks by death using a path-specific effects framework.^14^ This allows us to evaluate the number of CVD cases attributable to each metal through the longitudinal trajectory of CAC, and the number of CVD cases that do not happen due to participants dying during the follow-up.

## Methods

### Study Population

The MESA is a multi-center, prospective cohort study of subclinical and clinical CVD.^15^ Between 2000 and 2002, 6,814 participants were recruited at six study sites in Baltimore MD, Chicago IL, Los Angeles CA, New York NY, St. Paul MN, and Winston Salem NC. Participants were 45–84 years old, from four race and ethnic groups (White, Black, Hispanic and Chinese) and free of clinical CVD at baseline. They completed up to five clinic visits (2000-2002, 2002-2004, 2004-2006, 2005-2007, and 2010-2012). Informed consent was given by participants who met enrollment criteria. The Institutional Review Board at each study site approved the study.

The inclusion criteria for this study have been described elsewhere^11^ and are detailed in the Supplementary Methods. The final sample size included 6,527 participants with repeated measures of CAC.

### Urinary Metals

The analytical protocol to measure metals in MESA have been published.^16^ Briefly, urine samples were taken at MESA exam 1 (2,000-2,002) and metals were analyzed using Inductively Coupled Plasma Mass Spectrometry after dilution.^17^ Metal concentrations were divided by urine creatinine concentrations, measured using the Jaffe reaction method,^18^ to correct for urine dilution, and are expressed as µg metal/g creatinine. We included six metals with prior evidence of being associated with CAC progression and incident CVD in MESA:^11,12^ cadmium, cobalt, copper, uranium, tungsten, and zinc.

### Cardiac Computed Tomography (CT) Scanning and Coronary Artery Calcification

Procedures of cardiac CT scans to measure CAC in MESA have been described previously.^19^ Arterial trajectories across the surface of the heart were determined within 8 mm. Then, a phantom-based adjustment was applied, and candidate calcified plaques were identified. The criteria for calcification were for each plaque to be composed of at least 4 contiguous voxels with an attenuation level of 130 Hounsfield units or greater.^19^

Several methods exist to quantify CAC. The traditional Agatston score (AS) has the limitation that it ignores the presence of calcification below a certain threshold, classifying levels early in the calcification stages as CAC-AS=0.^20,21^ The spatially weighted calcium score (CAC-SW) is a semi-automated threshold-free CAC scoring method. This method assigns weights to each image voxel according to the phantom and neighboring voxels, thus maximizing the CT scan information. The detailed algorithm for calculation, providing the continuous CAC score, has been published.^20^ CAC-SW has shown to predict incident CHD events even among participants with CAC-AS=0.^20^ We used longitudinal measurements of CAC-SW across MESA exams 1-5 for the analyses. We used Multiple Imputation in Chained Equations (MICE) imputation^22^ to impute missing values in CAC in exams 1-5, based on the values of the other visits and other covariates. Details of the MICE procedure can be found in the Supplementary Methods and in Supplementary Table 1.

### Cardiovascular Disease Ascertainment

Participants were followed for incident cardiovascular events and mortality through December 2019; the median (interquartile range, IQR) follow up was 17.5 (11.2, 18.4) years. CVD diagnosis procedures in MESA have been published elsewhere^15^ and are detailed in the Supplementary Methods. Incident CVD events were defined as myocardial infarction, resuscitated cardiac arrest, definite and probable angina, stroke, atherosclerotic deaths including CHD and other CVD deaths. Information about death events was systematically collected from the National Death Index database.^23^ Participants were followed until the first CVD record of any type, death, loss to follow-up, or the administrative censoring date (December 31, 2019), whichever happened first.

### Covariates

Self reported age (years), sex (male, female), race and ethnicity (White, Black, Hispanic, or Chinese), education, or college degree or higher education), smoking status (never, former, current smoker), cigarette pack-years (calculated by multiplying the intensity in packs per day by duration in years) and use of lipid-lowering and hypertension medications (yes, no) were collected using questionnaires at MESA exam 1. Height and weight were measured to calculate body mass index (BMI, kg/m^2^). Resting systolic and diastolic blood pressure were measured 3 times in the seated position using a Dinamap model Pro 100 automated oscillometric sphygmomanometer with the last 2 measurements averaged for analysis. Low- and high-density lipoprotein cholesterol (LDL, HDL, mg/dL blood), and calibrated fasting plasma glucose (FPG, mg/dL plasma), were assessed using standard laboratory techniques. Diabetes mellitus (DM) was defined by the 2003 American Diabetes Association fasting criteria and categorized by normal (<100 mg/dL plasma FPG) or impaired fasting glucose (100-125 mg/dL plasma FPG), untreated or treated diabetes (≥126 mg/dL plasma FPG or taking diabetes medications). Estimated glomerular filtration rate (eGFR, mL/min/1.73 m^2^) is a measure of kidney function and can influence both metal excretion in urine and CVD risk. It was calculated using the new creatinine and cystatin-C based Chronic Kidney Disease Epidemiology Collaboration equation, without accounting for race and ethnicity.^24^

### Statistical Methods

Figure 1 depicts the directed acyclic graph for our setting. We performed a longitudinal mediation analysis using the CAC data collected at baseline and follow-up (exams 1-5). Our exposure variables were the six creatinine-corrected priority metal concentrations evaluated at exam 1, in separate models, and our outcome was time to first CVD event in years. We used an adaptation of the mediational g-formula that we developed for survival mediation analysis accounting for competing risks. We used a path-specific effects framework for a continuous exposure and a time-to-event outcome to conduct longitudinal mediation analysis accounting for competing risks^25^ (see Supplementary Methods). To do so, we consider death as another mediator which has a deterministic relationship with both the mediator of interest (CAC) and the outcome (in the sense that, if an individual dies, no further values of the mediator can be observed, and the outcome cannot happen).

**Figure 1.**
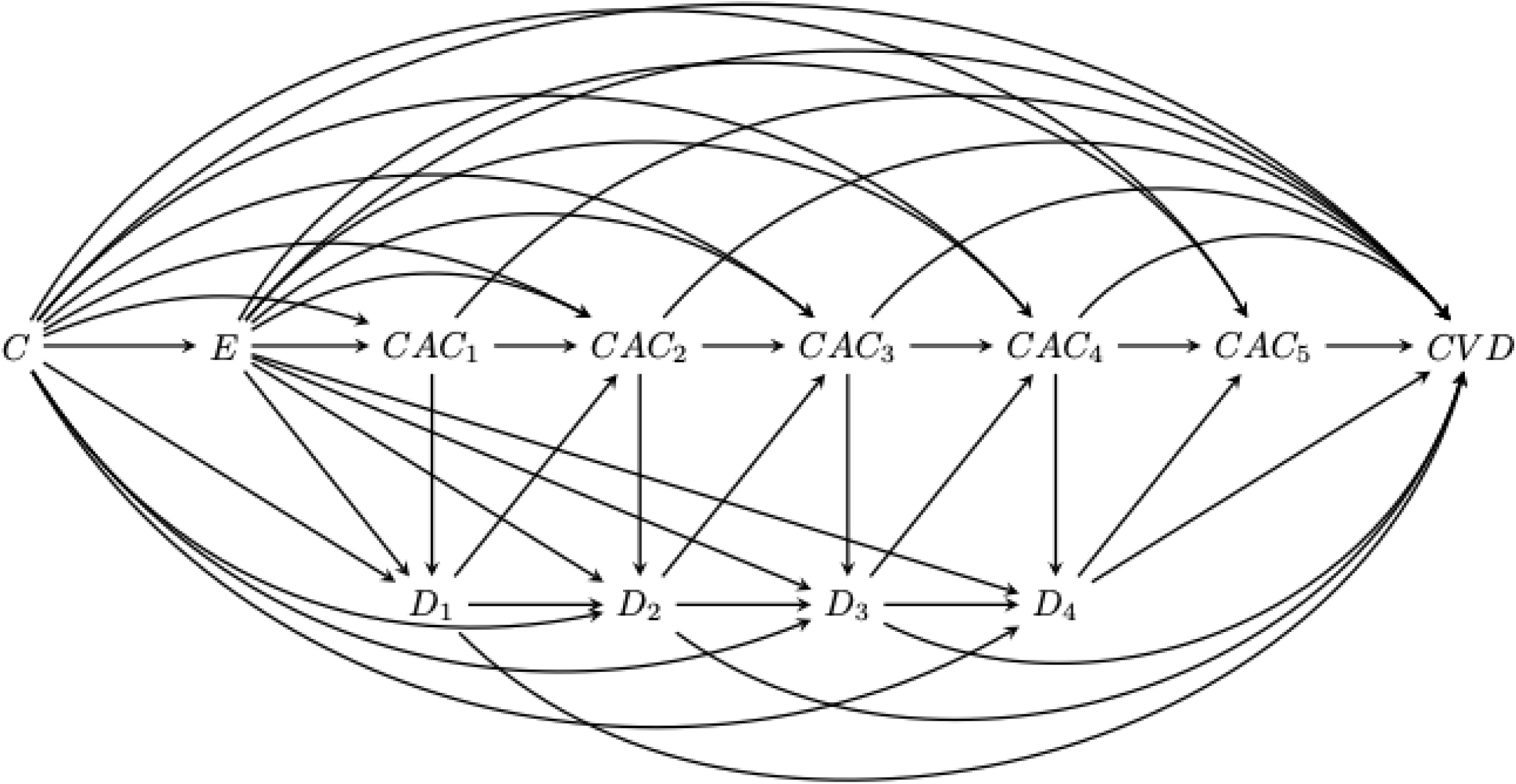
Directed Acyclic Graph of the association between each urinary metal (E) and time to cardiovascular disease (CVD) potentially mediated through the longitudinal trajectory of coronary artery calcification (CAC). Abbreviations: C, confounders; E, exposures (urinary metals); CAC_i_, coronary artery calcification measures at exam i; D_i_, death indicator at exam i; CVD, cardiovascular disease.

We used a semi-parametric additive hazards model with CVD incidence as the outcome and the metal of interest as the exposure, adjusted for CAC (in all visits) and baseline age, sex, race/ethnicity, study site, education, smoking status, cigarette pack-years, BMI, and eGFR (model 1). We further adjusted for clinical CVD risk factors (LDL and HDL cholesterol, diabetes mellitus, sBP, lipid lowering and hypertension medication, model 2), which could also be mediators on the association between metals and CVD. The code of the algorithm is publicly available and can be found in the following Github repository: https://github.com/arcedr/CM_CR_gFormula.

Our model reports the effects in differences in hazards (number of cases attributable to an IQR increase in the exposure per number of person-years^26^), which constitutes an absolute measure of risk that is relevant for public health. While differences in hazards are popular in epidemiological studies and represent a measure of impact for public health, recent work has demonstrated that they do not have a causal inference interpretation.^27^ Thus, in addition to differences in hazards, we provide results in differences in survival probability scale, i.e., being free of CVD after 20 years. Confidence intervals were calculated using quantile bootstrap, and were corrected by multiple comparisons using the Bonferroni approach to account for the fact that we are analyzing six metals in different models.

We ran the mediational g-formula algorithm with 1,000 iterations, comparing the metals’ 75^th^ to 25^th^ percentiles. We thus have four quantities of interest:

- *Total effect*: number of CVD cases per 100,000 person-years attributable to an IQR increase in each metal concentration.
- *Direct effect*: association of the metal with CVD through pathways not involving CAC or death. Number of CVD cases per 100,000 person-years attributable to an IQR increase in each metal concentration that do not happen through CAC or death pathways.
- *Indirect effect through CAC* (the mediator of interest): association between the metal and CVD through the CAC trajectory. Number of CVD cases per 100,000 person-years attributable to an IQR increase in each metal concentration that happen through the CAC trajectory.
- *Indirect effect through death* (the competing event): association between the metal and CVD through death (i.e., preventing CVD from happening). Number of CVD cases per 100,000 person-years attributable to an IQR increase in each metal concentration avoided by the fact that the individual died before the CVD event happened.

## Results

There were 1,140 incident CVD events, with mean follow-up time of 9 years for cases, and of 16 years for non-cases. Participants that developed CVD were older, more likely to be male, have treated diabetes mellitus, had lower education and eGFR levels, and had higher sBP levels (Table 1). CAC-SW increased over time and was higher among those who developed CVD. Median CAC-SW at baseline was 4.2 for those who did not have CVD, and 56.6 for those who had CVD. At exam 5, median CAC-SW was 60.1 for those who did not have CVD, and 253.6 for those who had CVD. Baseline urine levels of cadmium, cobalt, copper and zinc were higher among those who developed CVD.

**Table 1.** Participant characteristics by cardiovascular disease (CVD) status.

| Variables | No CVD (N=5,387) | Incident CVD (N=1,140) |
| --- | --- | --- |
| Age (years) | 61 (53, 69) | 68 (60, 75) |
| Sex % |  |  |
| Female | 55.1 | 42.2 |
| Male | 44.9 | 57.8 |
| Education level % |  |  |
| ≤ High School | 34.7 | 42.2 |
| Some College | 23.3 | 23.6 |
| > College | 42.0 | 34.2 |
| BMI (kg/m<sup>2</sup>) | 27.4 (24.4, 31.1) | 27.9 (25.1, 31.3) |
| eGFR (mL/min/1.73m<sup>2</sup>) | 78.4 (67.9, 89.4) | 73.5 (62.4, 85.7) |
| Smoking % |  |  |
| Never | 51.4 | 46.1 |
| Former | 35.8 | 40.9 |
| Current | 12.8 | 13.0 |
| Race / ethnicity % |  |  |
| White | 38.5 | 39.9 |
| Chinese | 12.3 | 10.3 |
| Black | 27.6 | 27.2 |
| Hispanic | 21.6 | 22.6 |
| LDL cholesterol mg/dL | 116 (96, 136) | 114 (95, 137) |
| HDL cholesterol mg/dL | 49 (41, 60) | 46 (39, 56) |
| Diabetes % |  |  |
| Normal | 76.4 | 63.6 |
| IFG | 13.3 | 15.0 |
| Untreated DM | 2.4 | 3.2 |
| Treated DM | 7.9 | 18.2 |
| SBP mmHg | 122 (110, 138) | 133 (118, 149) |
| Lipid medication % |  |  |
| No | 85.0 | 78.2 |
| Yes | 15.0 | 21.8 |
| Hypertension medication % |  |  |
| No | 65.8 | 49.5 |
| Yes | 34.2 | 50.5 |
| CAC-SW exam 1 | 4.2 (0.5, 35.4) | 56.6 (5.9, 237.9) |
| CAC-SW exam 2 | 7.1 (0.8, 48.6) | 69.2 (9.2, 294.0) |
| CAC-SW exam 3 | 54.8 (7.9, 256.0) | 296.6 (51.7, 1088.1) |
| CAC-SW exam 4 | 34.1 (6.5, 197.0) | 221.6 (35.5, 829.0) |
| CAC-SW exam 5 | 60.1 (10.8, 240.2) | 253.6 (55.4, 684.1) |
| Cadmium µg/g | 0.53 (0.35, 0.80) | 0.55 (0.36, 0.81) |
| Cobalt µg/g | 0.39 (0.28, 0.57) | 0.40 (0.29, 0.56) |
| Copper µg/g | 12.3 (10.0, 15.6) | 13.0 (10.4, 16.8) |
| Uranium µg/g | 0.005 (0.003, 0.011) | 0.005 (0.003, 0.011) |
| Tungsten µg/g | 0.06 (0.04, 0.10) | 0.06 (0.04, 0.11) |
| Zinc µg/g | 527 (356, 788) | 614 (413, 935) |
| Follow-up time (years) | 17.8 (14.3, 18.5) | 9.2 (4.4, 13.3) |

Abbreviations: CVD, cardiovascular disease; eGFR, estimated glomerular filtration rate; LDL, low density lipoprotein; HDL, high density lipoprotein; IFG, impaired fasting glucose; DM, diabetes mellitus; SBP, systolic blood pressure; CAC-SW, spatially weighted calcification score.
The table shows median (interquartile range) for continuous variables, percentages for categorical variables.

The total effects (attributable CVD cases/100,000 person-years (95% CI) per one IQR increase in metal levels) ranged from 88 (−83, 264) for uranium to 315 (168, 474) for copper (Table 2, Figure 2). The indirect effect for the association between metals and CVD through the CAC trajectory was statistically significant for all metals except cobalt and uranium in models not adjusted for clinical risk factors (model 1). The number of CVD cases/100,000 person-years (95% CI) attributable to an IQR increase in metal levels through the longitudinal trajectory of CAC (mediated effect) ranged from 19 (3, 38) for cobalt to 42 (24, 64) for zinc (Table 2, Figure 2). The indirect effect through death (avoided CVD cases/100,000 person-years (95% CI) due to death from causes other than CVD, i.e., CVD cases that did not occur due to death related to exposure to the metal) was −10 (−16, −4) for cadmium, and not statistically significant for the other metals. When evaluating the difference in probability of being free of CVD after 20 years, the indirect effect through CAC was statistically significant for all metals (Table 3, Figure 2). However, the indirect effect through death was only statistically significant for cadmium.

**Figure 2.**
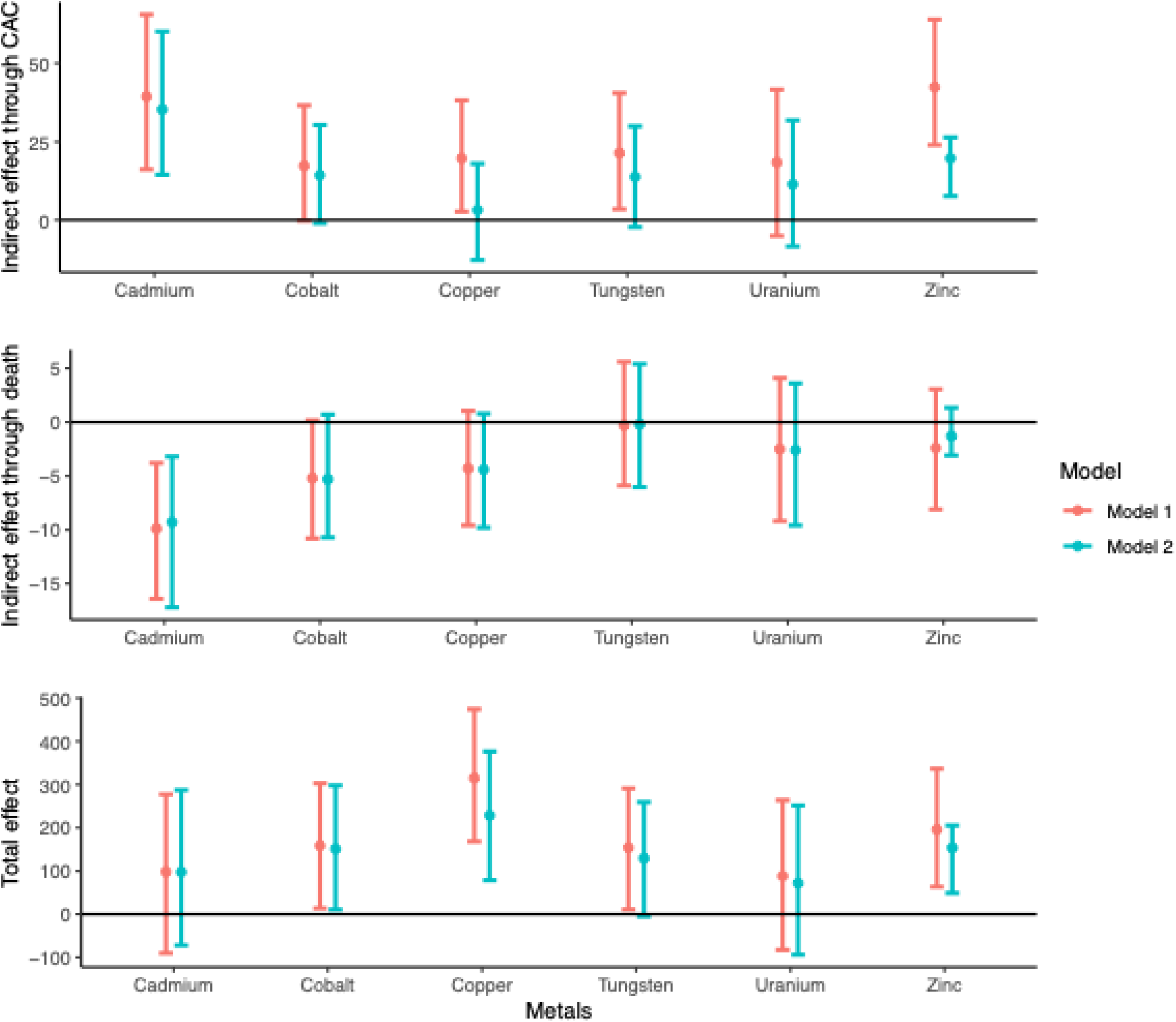
Direct, indirect and total effects (number of cardiovascular disease cases per 100,000 person-years) attributable to metals through the spatially weighted calcium scores and death trajectories. The y axis represents number of cardiovascular disease (CVD) cases attributable to an interquartile range increase in each metal per 100,000 person-years. For the indirect effect through death, the numbers represent number of CVD cases attributable to an interquartile range increase in each metal avoided due to death per 100,000 person-years. Model 1: adjusted for age, sex, race/ethnicity, study site, education level, body mass index, estimated glomerular filtration rate smoking status, and cigarette pack-years. Model 2: additionally adjusted for LDL cholesterol, HDL cholesterol, sBP, diabetes status, lipid lowering medication and hypertension medication.

**Table 2.**
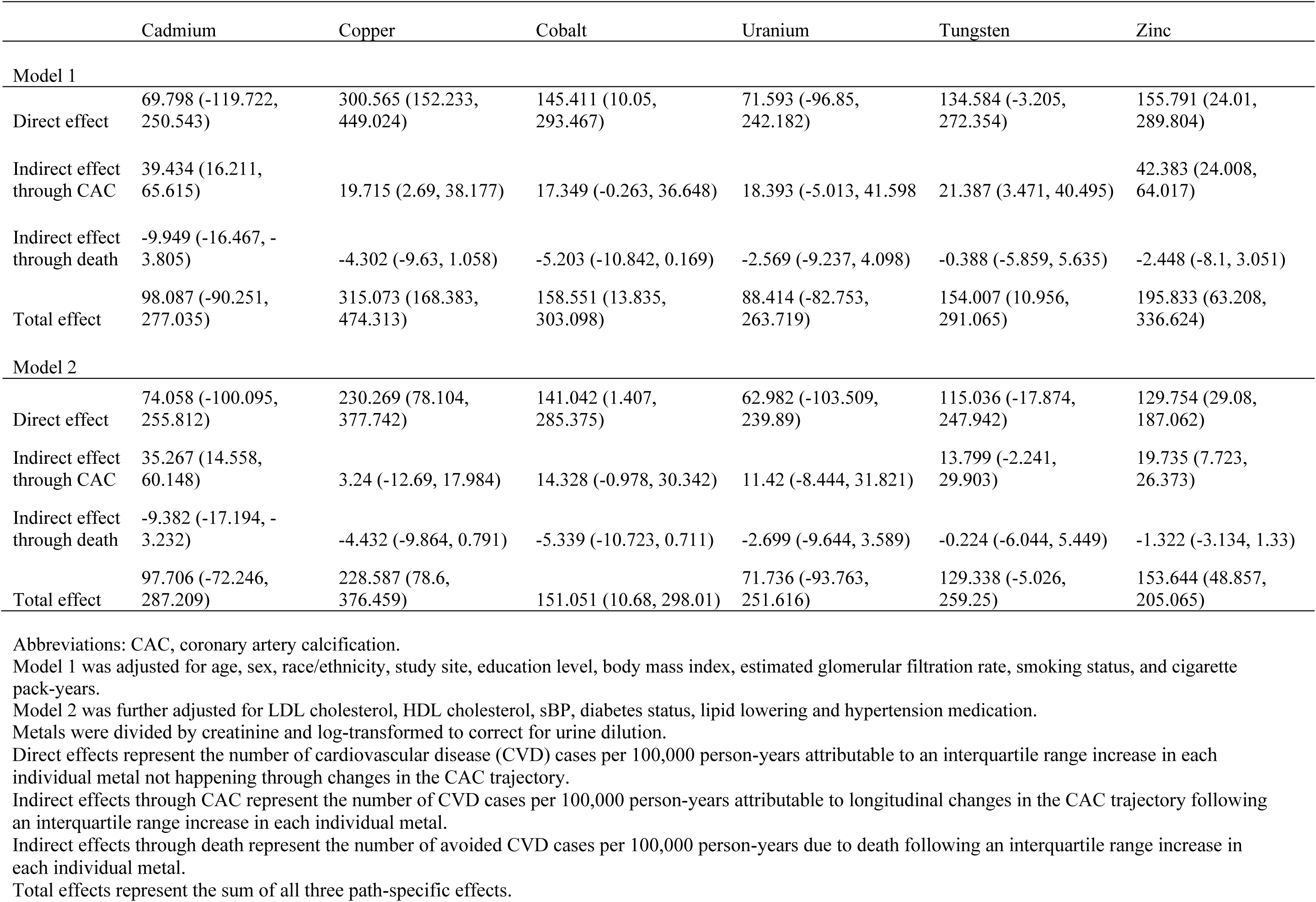
Direct, indirect and total effects (number of cardiovascular disease cases per 100,000 person-years) of metals on cardiovascular disease incidence through longitudinal changes in spatially weighted calcium scores accounting for competing risks by death in the differences in hazards scale.

|  | Cadmium | Copper | Cobalt | Uranium | Tungsten | Zinc |
| --- | --- | --- | --- | --- | --- | --- |
| **Model 1** |  |  |  |  |  |  |
| Direct effect | 69.798 (-119.722, 250.543) | 300.565 (152.233, 449.024) | 145.411 (10.05, 293.467) | 71.593 (-96.85, 242.182) | 134.584 (-3.205, 272.354) | 155.791 (24.01, 289.804) |
| Indirect effect through CAC | 39.434 (16.211, 65.615) | 19.715 (2.69, 38.177) | 17.349 (-0.263, 36.648) | 18.393 (-5.013, 41.598) | 21.387 (3.471, 40.495) | 42.383 (24.008, 64.017) |
| Indirect effect through death | -9.949 (-16.467, -3.805) | -4.302 (-9.63, 1.058) | -5.203 (-10.842, 0.169) | -2.569 (-9.237, 4.098) | -0.388 (-5.859, 5.635) | -2.448 (-8.1, 3.051) |
| Total effect | 98.087 (-90.251, 277.035) | 315.073 (168.383, 474.313) | 158.551 (13.835, 303.098) | 88.414 (-82.753, 263.719) | 154.007 (10.956, 291.065) | 195.833 (63.208, 336.624) |
| **Model 2** |  |  |  |  |  |  |
| Direct effect | 74.058 (-100.095, 255.812) | 230.269 (78.104, 377.742) | 141.042 (1.407, 285.375) | 62.982 (-103.509, 239.89) | 115.036 (-17.874, 247.942) | 129.754 (29.08, 187.062) |
| Indirect effect through CAC | 35.267 (14.558, 60.148) | 3.24 (-12.69, 17.984) | 14.328 (-0.978, 30.342) | 11.42 (-8.444, 31.821) | 13.799 (-2.241, 29.903) | 19.735 (7.723, 26.373) |
| Indirect effect through death | -9.382 (-17.194, -3.232) | -4.432 (-9.864, 0.791) | -5.339 (-10.723, 0.711) | -2.699 (-9.644, 3.589) | -0.224 (-6.044, 5.449) | -1.322 (-3.134, 1.33) |
| Total effect | 97.706 (-72.246, 287.209) | 228.587 (78.6, 376.459) | 151.051 (10.68, 298.01) | 71.736 (-93.763, 251.616) | 129.338 (-5.026, 259.25) | 153.644 (48.857, 205.065) |

Abbreviations: CAC, coronary artery calcification.
Model 1 was adjusted for age, sex, race/ethnicity, study site, education level, body mass index, estimated glomerular filtration rate, smoking status, and cigarette pack-years.
Model 2 was further adjusted for LDL cholesterol, HDL cholesterol, sBP, diabetes status, lipid lowering and hypertension medication.
Metals were divided by creatinine and log-transformed to correct for urine dilution.
Direct effects represent the number of cardiovascular disease (CVD) cases per 100,000 person-years attributable to an interquartile range increase in each individual metal not happening through changes in the CAC trajectory.
Indirect effects through CAC represent the number of CVD cases per 100,000 person-years attributable to longitudinal changes in the CAC trajectory following an interquartile range increase in each individual metal.
Indirect effects through death represent the number of avoided CVD cases per 100,000 person-years due to death following an interquartile range increase in each individual metal.
Total effects represent the sum of all three path-specific effects.

**Table 3.** Direct, indirect and total effects (number of cardiovascular disease cases per 100,000 person-years) of metals on cardiovascular disease incidence through longitudinal changes in spatially weighted calcium scores accounting for competing risks by death in the differences in survival probability scale.

|  | Cadmium | Copper | Cobalt | Uranium | Tungsten | Zinc |
| --- | --- | --- | --- | --- | --- | --- |
| **Model 1** |  |  |  |  |  |  |
| Direct effect (%) | -1.022 (-3.016, 1.072) | -4.592 (-6.199, -2.941) | -2.223 (-3.638, -0.646) | -1.079 (-3.039, 0.873) | -2.097 (-3.557, -0.589) | -2.396 (-3.758, -1.111) |
| Indirect effect through CAC (%) | -0.607 (-0.937, -0.315) | -0.294 (-0.494, -0.106) | -0.285 (-0.483, -0.08) | -0.288 (-0.543, -0.051) | -0.238 (-0.443, -0.043) | -0.635 (-0.855, -0.421) |
| Indirect effect through death (%) | 0.138 (0.056, 0.227) | 0.061 (-0.007, 0.127) | 0.074 (-0.003, 0.149) | 0.037 (-0.053, 0.126) | 0.004 (-0.072, 0.08) | 0.036 (-0.04, 0.113) |
| Total effect (%) | -1.49 (-3.533, 0.736) | -4.84 (-6.451, -3.163) | -2.438 (-3.877, -0.82) | -1.315 (-3.277, 0.651) | -2.334 (-3.803, -0.787) | -2.991 (-4.419, -1.696) |
| **Model 2** |  |  |  |  |  |  |
| Direct effect (%) | -1.309 (-3.26, 0.717) | -3.47 (-5.068, -2.08) | -2.112 (-3.602, -0.588) | -1.011 (-3.039, 0.827) | -1.838 (-3.368, -0.302) | -1.148 (-2.657, 0.27) |
| Indirect effect through CAC (%) | -0.567 (-0.826, -0.337) | -0.066 (-0.225, 0.086) | -0.249 (-0.429, -0.074) | -0.152 (-0.365, 0.068) | -0.155 (-0.317, 0.011) | -0.311 (-0.497, -0.126) |
| Indirect effect through death (%) | 0.135 (0.05, 0.232) | 0.061 (-0.004, 0.13) | 0.073 (0.001, 0.153) | 0.041 (-0.053, 0.13) | 0.003 (-0.078, 0.084) | 0.034 (-0.058, 0.104) |
| Total effect (%) | -1.729 (-3.681, 0.382) | -3.467 (-5.082, -2.034) | -2.29 (-3.799, -0.784) | -1.133 (-3.159, 0.685) | -1.994 (-3.54, -0.421) | -1.435 (-2.941, 0.004) |

Abbreviations: CAC, coronary artery calcification.
Model 1: adjusted for age, sex, race/ethnicity, study site, education level, body mass index, estimated glomerular filtration rate, smoking status, and cigarette pack-years.
Model 2: additionally adjusted for LDL cholesterol, HDL cholesterol, sBP, diabetes status, lipid lowering medication and hypertension medication.
Metals were log-transformed and divided by creatinine to correct for urine dilution.
Direct effects represent the difference in probability of being free of cardiovascular disease (CVD) after 20 years attributable to an interquartile range increase in each individual metal not happening through changes in the CAC trajectory.
Indirect effects through CAC represent the difference in probability of being free of CVD after 20 years attributable to longitudinal changes in the CAC trajectory following an interquartile range increase in each individual metal.
Indirect effects through death represent the difference in probability of being free of CVD after 20 years attributable to death following an interquartile range increase in each individual metal.
Total effects represent the sum of all three path-specific effects.

The associations were slightly attenuated when adjusting the models for clinical CVD risk factors, which could be related to clinical risk factors other than CAC serving as mediators on the association between metals and CVD. However, the indirect effect through CAC was still statistically significant for cadmium and zinc in the difference in hazards scale (Table 2, model 2, Figure 2), and for cadmium, cobalt, and zinc in the survival probability difference scale (Table 3, model 2, Figure 2), supporting an effect of metals on atherosclerosis progression independent of clinical risk factors.

## Discussion

This large, multi-ethnic, longitudinal study of adults supports that part of the association between increasing levels of essential and non-essential metals measured in urine and CVD is attributable to changes in CAC, a subclinical atherosclerosis marker, over time. Our novel statistical method allowed us to disentangle different pathways that contribute to the association between metals and CVD, incorporating a continuous mediator for repeated CAC-SW over time. For urinary cadmium, half of the association with incident CVD could be mediated by longitudinal changes in CAC. For copper and uranium, other pathways beyond CAC might be more relevant. Further, our approach enabled us to account for competing risks due to death when analyzing time to CVD events, which was particularly important for metals that are strongly associated with all-cause mortality, such as cadmium.

The importance of exposure to metals for CVD development has been extensively documented and recently acknowledged by the American Heart Association.^5^ Given the burden of CVD in the general US population, strategies for early detection and prevention are needed. The finding that part of the association of metals with CVD can be explained through changes in CAC––which occur years before the onset of clinical events––supports the upstream role of metal exposure and/or urinary excretion in CVD pathogenesis, and presents novel opportunities for early screening and prevention, as exposure to metals is, at least partially, modifiable.

Sources of exposure to metals might include individual-level sources and lifestyle factors such as smoking, e-cigarettes, dietary preferences, and consumer products, but also community level sources such as air, drinking water, or soil pollution.^28^ Previous evidence has documented declines in CVD risk after declines in exposure to toxic metals such as lead and cadmium through regulatory changes,^29,30^ revealing the potential of interventions to reduce exposure and improve related health effects. While interventions to remove metals accumulated in the body have not yet been successful in reducing clinical CVD events, the non-significant inverse association between disodium edetate, an active chelating agent that removes divalent cations (e.g., lead) from the body, compared to placebo (HR 0.93 [0.76, 1.16]) in the Trial to Assess Chelation Therapy 2 (TACT2),^31^ suggests a public health benefit for interventions that prevent metal exposures. Yet, assessment of metal removal treatment on subclinical CVD has not been studied.

Evidence has consistently shown that exposure to cadmium is a risk factor for CVD.^3,32^ Prior work in MESA showed that urinary cadmium has a strong association with CAC progression.^11^ Other cross-sectional studies in populations from Sweden^33^ and Spain^34^ have shown consistent findings. Our findings indicate that a large portion of the effect of cadmium on CVD is mediated through changes in CAC. Cadmium also showed the highest indirect effect through death, which is an indicator of strong competing risks by death, and thus, of the strong association of cadmium with death for causes other than CVD. This is consistent with previous studies reporting strong associations between cadmium and all-cause mortality in MESA and other cohorts,^12,35–37^ and highlights the importance of accounting for mortality due to other diseases associated with cadmium, such and cancer ^38^ or lung disease.^39^

Essential metals are needed in the body for cellular growth and development.^40^ However, increasing levels of essential metals in extracellular compartments such as serum or urine may not reflect excess intake, but early cardiometabolic dysregulation and release from the intracellular compartment.^41,42^ The fact that essential metals such as copper and zinc showed the strongest association with CVD is remarkable, and while it has been reported in several populations,^43^ it remains a largely overlooked concept. Our mediation analysis shows that most of the association between urinary copper and CVD might involve other alternative pathways beyond CAC. Other pathways such as diabetes, which has been associated with elevated urinary zinc and copper levels,^44,45^ might be playing a key role in these associations. Future work in the statistical methods development field should include methods for the evaluation of multiple longitudinal mediating pathways simultaneously, in order to shed light on the different pathways that are involved in metal-related CVD.

Our novel statistical method allowed us to disentangle the different pathways that contribute to the association between metals and CVD, incorporating a continuous mediator that leverages the CAC-SW for measuring CAC status, and to evaluate the role of competing risks, specifically, deaths from non-CVD causes. Traditional Cox proportional hazards models censor people that die as if they had dropped out of the study, and therefore consider they could develop the disease at the same rate as those who remain in the study, leading to a potential overestimation or underestimation of the association. Other approaches for competing risks in mediation analysis include the survivor average causal effect^46^ and separable effects.^47^ These methods, however, are focused on a population that is either unrealistic (all survivors), or not observed (those who would survive no matter the exposure level). Our method decomposes the contribution of each pathway to the association between an exposure and a health outcome while accounting for people that die during follow-up. This provides a better opportunity to investigate different pathways involved in the adverse health effects of elevated urinary metal levels. Other strengths include the large sample size and long follow-up, the inclusion of ethnically diverse participants from different geographic locations, the high-quality surveillance of CVD outcomes, the use of the SWCS, a more sensible and accurate method than the traditional Agatston score, to measure CAC, and the ability to account for a wide range of confounders.

Our study has several limitations. First, we rely on a single time point for urinary metals measurement. Previous work in MESA using a subset of participants with metals measured in exam 1 and 5 (8-11 years apart) have shown mostly consistent results,^11^ supporting that single urinary metal measures might reflect long-term exposure and internal dose. Our g-formula algorithm considers discrete time points for death, rather than modeling death as time-to-event, which leads to a loss of granularity and potential measurement error. Future work should include the extension of this algorithm to multi-state models,^48^ in which time to death and time to event of interest are considered jointly. In addition, we used MICE to imputed missing SWCS values as our model does not currently support incomplete longitudinal data. Future work should focus on an extension of this method to mixed effects models, in order to compare imputed results to those fitted in incomplete data. Overall, caution is needed when interpreting these results because as in any observational study, unmeasured confounding cannot be ruled out.

In conclusion, our method allowed us to identify that the association of urinary cadmium, copper, cobalt, uranium, tungsten, and zinc with CVD might be partially mediated by longitudinal changes in CAC. This pathway might be particularly important for cadmium, cobalt, zinc, and tungsten, while other biological pathways might be more relevant for copper. Additionally, we were able to simultaneously characterize, for the first time, the pathway through mortality, which explains to what extent increases in metal concentrations are associated with a reduction in CVD cases due to an increase in competing events that might be more lethal than CVD. Our results could inform strategies for early detection and prevention of CVD based on CAC measurements, and the implementation of preventive strategies to address CVD risk by reducing exposure to metals, especially for cadmium.

## Data Availability

Data can be available to investigators upon request through the Multi-Ethnic Study of Atherosclerosis website: https://www.mesa-nhlbi.org/

https://github.com/arcedr/CM_CR_gFormula

## Acknowledgements

The authors thank the investigators, the staff, and the participants of the MESA study for their valuable contributions. A full list of participating MESA investigators and institutions can be found at http://www.mesa-nhlbi.org. This paper has been reviewed and approved by the MESA Publications and Presentations Committee. The funder had no role in the design and conduct of the study; collection, management, analysis, and interpretation of the data; preparation, review, or approval of the manuscript; and decision to submit the manuscript for publication.

